# Clinical characteristics of 36 non-survivors with COVID-19 in Wuhan, China

**DOI:** 10.1101/2020.02.27.20029009

**Authors:** Ying Huang, Rui Yang, Ying Xu, Ping Gong

**Affiliations:** Department of Oncology, the Fifth Hospital of Wuhan, Wuhan, China; Department of Vascular surgery, the Fifth Hospital of Wuhan, Wuhan, China; Department of Endocrinology, the Fifth Hospital of Wuhan, Wuhan, China; Physical Examination Center, the Fifth Hospital of Wuhan, Wuhan, China

## Abstract

**Background:** Although the outbreak of Coronavirus disease 2019 (COVID-19) has caused over 2200 deaths in China, there was no study about death yet. We aimed to describe the clinical characteristics of non-survivors with COVID-19.

**Methods:** For this retrospective, single-center study, we included 36 non-survivors with COVID-19 in the Fifth Hospital of Wuhan. Cases were confirmed by real-time RT-PCR between Jan 21 and Feb 10, 2020 according to the recommended protocol. The epidemiological, demographic, clinical, laboratory, radiological and treatment data were collected and analyzed. Outcomes were followed up until Feb 14, 2020. This study was approved by the ethics commissions of the Fifth Hospital of Wuhan, with a waiver of informed consent due to a public health outbreak investigation.

**Findings:** We included 36 patients who died from COVID-19. The mean age of the patients was 69.22 years (SD 9.64, range 50-90). 25(69.44%) patients were males, and 11 (30.56%) female. 26 (72.22%) patients had chronic diseases, mainly including hypertension, cardiovascular disease and diabetes. Patients had common clinical symptoms of fever (34 [94.44%] patients), cough (28 [77.78%] patients), shortness of breath (21 [58.33%] patients), and fatigue (17 [47.22%] patient). Chest computed tomographic scans showed that 31 (96.88%) patients had bilateral pneumonia. Lymphopenia (lymphocyte count, 0.67□×□109/L [SD, 0.33]) occurred in 24 patients (70.59%), decreased albumin (30.18, [SD, 4.76]) in 25 patients (80.65%), elevated D-dimer (8.64 [IQR, 2.39-20]) in 27 patients (100%), and elevated lactate dehydrogenase (502.5 U/L [IQR, 410-629]) in 26 patients (100%). Nearly all of the patients have elevated CRP (106.3 mg/L [IQR, 60.83-225.3]), PCT (0.61 ng/ml [IQR, 0.16-2.10]) and IL-6 (100.6 pg/ml [IQR, 51.51-919.5]). Most patients received antiviral therapy and antibiotic therapy, and more than half of patients received glucocorticoid therapy (25 [69.44%]). All the patients had acute respiratory distress syndrome (ARDS). The median time from onset to ARDS was 11 days. One (2.78%) patient presented with acute renal injury. The median time from onset to death was 17 days.

**Interpretation:** Lots of patients died from COVID-19 till now. The median survival time of these non-survivors from onset to death was about 2 weeks. Most patients were older males with comorbidities. They finally progressed to ARDS. The median time from onset to ARDS was 11 days. Gradually decreased lymphocytes and increased inflammation biomarkers were common, and need to be monitored in the routine treatment.

**Funding:** There is no any funder involved in this study.

**Research in context:** *Evidence before this study:* SARS-CoV-2 has been spreading in China as well as in other countries. We searched PubMed for articles published up to Feb 15, 2020. Serveral articles that describe the epidemiological and clinical characteristics of general COVID-19 patients. However, special reports about dead cases of COVID-19 were limited but warranted, considering the large amount of confirmed cases, which is still increasing

*Added value of this study:* We retrospectively analysed specific clinical information of 36 non-survivors infected with SARS-CoV-2 in this single-centered study. Most patients were older males with comorbidities. Gradually decreased lymphocytes and increased inflammation biomarkers were found in these patients. They finally progressed to ARDS. The median time from onset to ARDS was 11 days. The mean survival time in our cohort of COVID-19 non-survivors was about 2 weeks.

*Implications of all the available evidence:* Lots of patients died from COVID-19 till now. The median survival time of these non-survivors from onset to death was about 2 weeks. Most patients were older males with comorbidities. They finally progressed to ARDS. Early detection and intervention of patients are especially important which can delay the development from mild to severe cases.

## Introduction

The outbreak of Coronavirus disease 2019 (COVID-19) caused by severe acute respiratory syndrome coronavirus 2 (SARS-CoV-2) happened since December 2020 in Wuhan^1^. ^2^ SARS-CoV-2 has been spreading in China as well as in other countries^3–6^. The outbreak of SARS-CoV-2 infection has been declared as a Public Health Emergency of International Concern by the World Health Organization (WHO) on 30 January 2020. According to the situation report from WHO, as of 21 February 2020, SARS-Cov-2 infection has caused 76769 confirmed cases in 27 countries and 2247 deaths globally^7^.

Severity of COVID-19 varied from mild to severe.^8^ Mortality rate of COVID-19 can be reduced, by timely identifying those who are at higher risk of developing into critically ill patients, closely monitoring changes of their disease course, and applying intensive care treatment to these patients. However, reports about dead cases of COVID-19 were limited but warranted, considering the large amount of confirmed cases, which is still increasing. In this single-centered study, we retrospectively analysed specific clinical information of 36 non-survivors infected with SARS-CoV-2.

## Methods

### Study design and participants

For this retrospective, single-center study, we included 36 non-survivors with COVID-19 in the Fifth Hospital of Wuhan. Cases were confirmed between Jan 21 and Feb 10, 2020 according to the recommended protocol. This study was approved by the ethics commissions of hospital, with a waiver of informed consent due to a public health outbreak investigation.

### Procedures

COVID-19 was confirmed by real-time RT-PCR using the same protocol described previously^9^. Additionally, all patients were given chest computed tomography (CT). We obtained epidemiological, demographic, clinical, laboratory, treatment and outcome data from patients’ medical records. Clinical outcomes were followed up to Feb 14, 2020.

### Outcomes

We analysed epidemiological, demographics, exposure history, smoking history, chronic medical illness, signs and symptoms on admission, comorbidity, chest CT findings, laboratory findings and treatment measures.

### Statistical analysis

Categorical variables were described as count (%). Continuous variables were described using mean (SD) if they are normally distributed, median (IQR) if they are not. All statistical analyses were performed using R (version 3.6.1).

## Results

We included 36 patients who died from COVID-19 in the Fifth Hospital of Wuhan. One (2.78%) patients had a history of exposure to the Huanan seafood market in Wuhan. The mean age of the patients was 69.22 years (SD 9.64, range 50-90). Twenty-five (69.44%) patients were males, and 11 (30.56%) female. Four (11.11%) patients have smoking history. Twenty-six (72.22%) patients had chronic diseases. The most common chronic diseases were hypertension (21, 58.33%), cardiovascular (8, 22.22%) and diabetes (7, 19.44%) (Table 1).

**Table 1:**
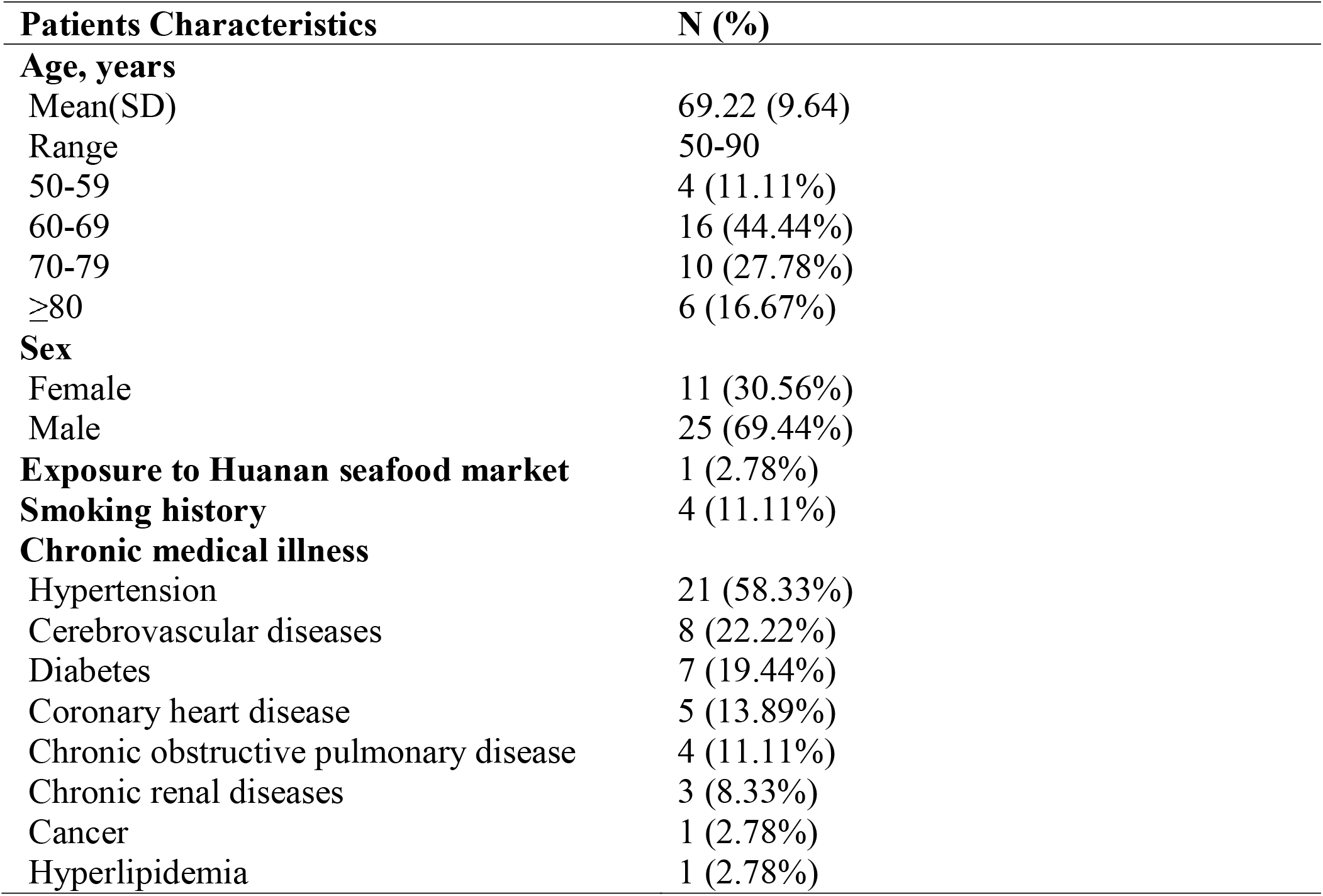
Baseline characteristics of 36 non-survivors admitted to the Fifth Hospital of Wuhan (Jan 23–Feb 10, 2020) with COVID-19

| **Patients Characteristics** | **N (%)** |
| --- | --- |
| **Age, years** |  |
| Mean(SD) | 69.22 (9.64) |
| Range | 50-90 |
| 50-59 | 4 (11.11%) |
| 60-69 | 16 (44.44%) |
| 70-79 | 10 (27.78%) |
| ≥80 | 6 (16.67%) |
| **Sex** |  |
| Female | 11 (30.56%) |
| Male | 25 (69.44%) |
| **Exposure to Huanan seafood market** | 1 (2.78%) |
| **Smoking history** | 4 (11.11%) |
| **Chronic medical illness** |  |
| Hypertension | 21 (58.33%) |
| Cerebrovascular diseases | 8 (22.22%) |
| Diabetes | 7 (19.44%) |
| Coronary heart disease | 5 (13.89%) |
| Chronic obstructive pulmonary disease | 4 (11.11%) |
| Chronic renal diseases | 3 (8.33%) |
| Cancer | 1 (2.78%) |
| Hyperlipidemia | 1 (2.78%) |

On admission, most patients had fever, cough and more than half of them had shortness of breath (Table 2). Other symptoms included fatigue, dyspnea, sputum production, disturbance of consciousness, diarrhea, hemoptysis and myalgia. (Table 2). Patients had common clinical symptoms of fever (34, 94.44%), cough (28, 38.89%), shortness of breath (21, 58.33%), fatigue (17, 47.22%), dyspnea (14, 38.89%), sputum production (8, 22.22%), disturbance of consciousness (8, 22.22%), diarrhea (3, 8.33%), hemoptysis (2, 5.56%), and muscle ache (1, 2.78%).

**Table 2:** Clinical characteristics, treatment and clinical outcomes of 36 non-survivors admitted to the Fifth Hospital of Wuhan (Jan 23–Feb 10, 2020) with COVID-19

| **Characteristics** | **N (%)** |
| --- | --- |
| **Signs and symptoms at admission** |  |
| Fever | 34 (94.44%) |
| Cough | 28 (77.78%) |
| Short of breath | 21 (58.33%) |
| Fatigue | 17 (47.22%) |
| Dyspnea | 14 (38.89%) |
| Sputum production | 8 (22.22%) |
| Disturbance of consciousness | 8 (22.22%) |
| Diarrhea | 3 (8.33%) |
| Hemoptysis | 2 (5.56%) |
| Myalgia | 1 (2.78%) |
| **Chest CT findings (N=32)** |  |
| Bilateral pneumonia | 31 (96.88%) |
| **Treatment** |  |
| Oxygen therapy | 35 (97.22%) |
| Mechanical ventilation |  |
| Non-invasive | 19 (52.78%) |
| Invasive | 9 (25%) |
| Antibiotic treatment | 36 (100%) |
| Antiviral treatment | 35 (97.22%) |
| Glucocorticoids | 25 (69.44%) |
| Intravenous immunoglobulin therapy | 20 (55.56%) |
| α-IFN | 6 (16.67%) |
| **Comorbid conditions** |  |
| ARDS | 36 (100%) |
| Electrolyte disturbance | 16 (44.44%) |
| Acute renal injury | 1 (2.78%) |
| **Clinical Outcome** |  |
| Median Time from onset to ARDS | 11 |
| Median Time from onset to death | 17 |

According to chest CT scan, 31 (96.88%, N=32) patients showed bilateral pneumonia (75%). On admission, leucocytes were below the normal range in 5 (14.71%) patients and above the normal range in 11 (32.35%) patients (Table 3). Neutrophils showed above the normal range in 16 (47.06%) patients. Lymphocytes and hemoglobin were below the normal range in many patients (Table 3). Twelve (35.39%) patients’ platelets were below the normal range. 22 patients had varying degrees of liver function abnormality, with alanine aminotransferase (ALT) or aspartate aminotransferase (AST) rose above the normal range (Table 3). Elevation of lactate dehydrogenase were showed in 26 (100%, N=26) patients. About 30% of these patients had different degrees of renal function damage, with elevated blood urea nitrogen or serum creatinine.

**Table 3:**
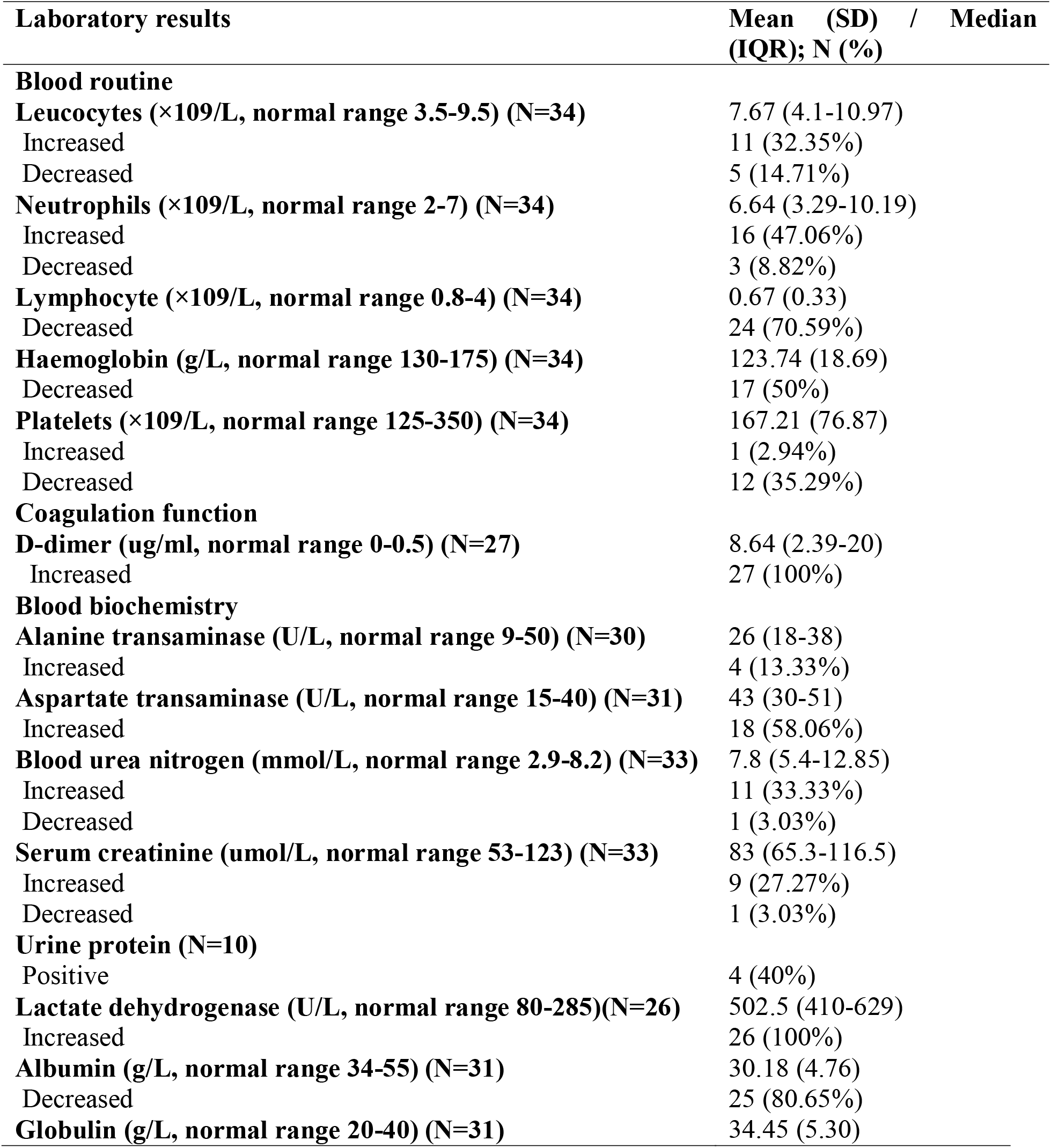
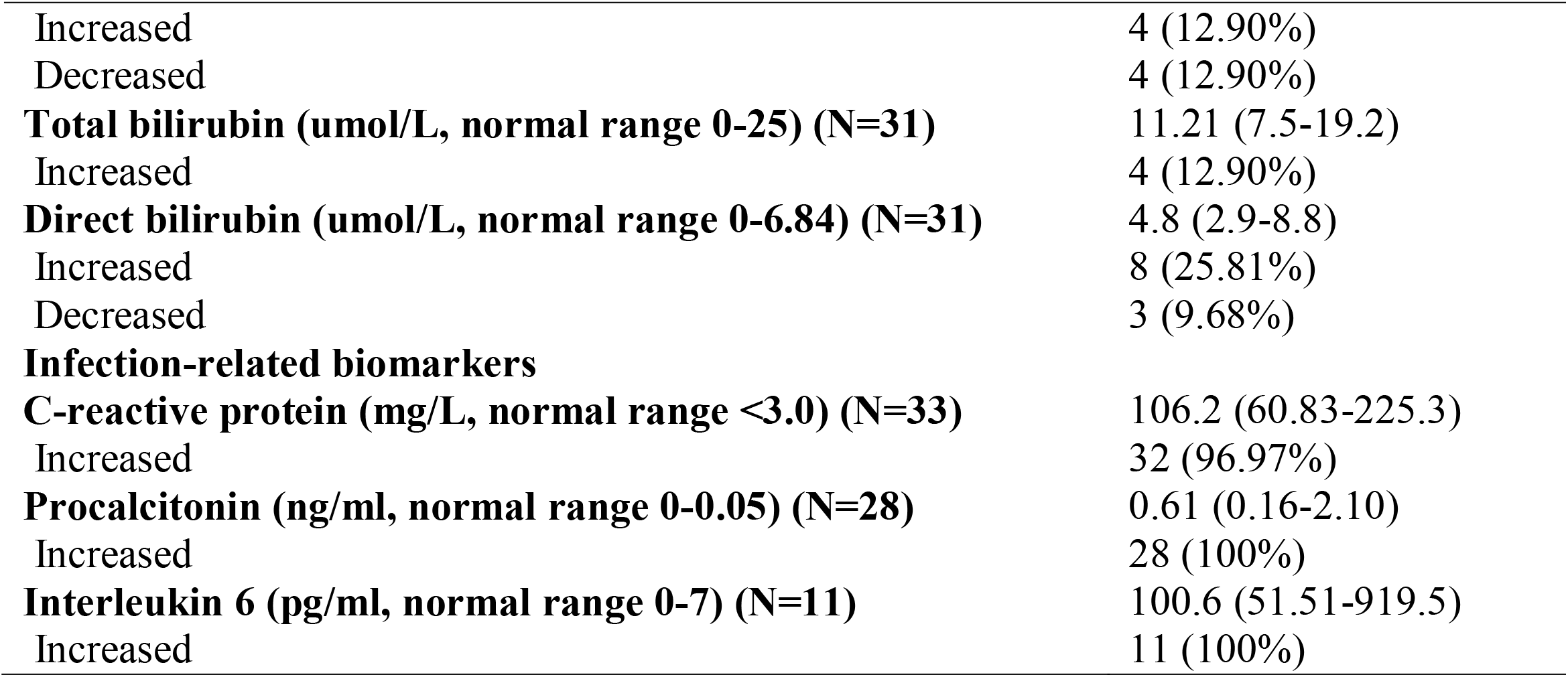
Laboratory results of non-survivors with COVID-19

| **Laboratory results** | **Mean (SD) / Median (IQR); N (%)** |
| --- | --- |
| **Blood routine** |  |
| **Leucocytes (×10<sup>9</sup>/L, normal range 3.5-9.5) (N=34)** | 7.67 (4.1-10.97) |
| Increased | 11 (32.35%) |
| Decreased | 5 (14.71%) |
| **Neutrophils (×10<sup>9</sup>/L, normal range 2-7) (N=34)** | 6.64 (3.29-10.19) |
| Increased | 16 (47.06%) |
| Decreased | 3 (8.82%) |
| **Lymphocyte (×10<sup>9</sup>/L, normal range 0.8-4) (N=34)** | 0.67 (0.33) |
| Decreased | 24 (70.59%) |
| **Haemoglobin (g/L, normal range 130-175) (N=34)** | 123.74 (18.69) |
| Decreased | 17 (50%) |
| **Platelets (×10<sup>9</sup>/L, normal range 125-350) (N=34)** | 167.21 (76.87) |
| Increased | 1 (2.94%) |
| Decreased | 12 (35.29%) |
| **Coagulation function** |  |
| **D-dimer (ug/ml, normal range 0-0.5) (N=27)** | 8.64 (2.39-20) |
| Increased | 27 (100%) |
| **Blood biochemistry** |  |
| **Alanine transaminase (U/L, normal range 9-50) (N=30)** | 26 (18-38) |
| Increased | 4 (13.33%) |
| **Aspartate transaminase (U/L, normal range 15-40) (N=31)** | 43 (30-51) |
| Increased | 18 (58.06%) |
| **Blood urea nitrogen (mmol/L, normal range 2.9-8.2) (N=33)** | 7.8 (5.4-12.85) |
| Increased | 11 (33.33%) |
| Decreased | 1 (3.03%) |
| **Serum creatinine (umol/L, normal range 53-123) (N=33)** | 83 (65.3-116.5) |
| Increased | 9 (27.27%) |
| Decreased | 1 (3.03%) |
| **Urine protein (N=10)** |  |
| Positive | 4 (40%) |
| **Lactate dehydrogenase (U/L, normal range 80-285)(N=26)** | 502.5 (410-629) |
| Increased | 26 (100%) |
| **Albumin (g/L, normal range 34-55) (N=31)** | 30.18 (4.76) |
| Decreased | 25 (80.65%) |
| **Globulin (g/L, normal range 20-40) (N=31)** | 34.45 (5.30) |
| Increased | 4 (12.90%) |
| Decreased | 4 (12.90%) |
| **Total bilirubin (umol/L, normal range 0-25) (N=31)** | 11.21 (7.5-19.2) |
| Increased | 4 (12.90%) |
| **Direct bilirubin (umol/L, normal range 0-6.84) (N=31)** | 4.8 (2.9-8.8) |
| Increased | 8 (25.81%) |
| Decreased | 3 (9.68%) |
| **Infection-related biomarkers** |  |
| **C-reactive protein (mg/L, normal range <3.0) (N=33)** | 106.2 (60.83-225.3) |
| Increased | 32 (96.97%) |
| **Procalcitonin (ng/ml, normal range 0-0.05) (N=28)** | 0.61 (0.16-2.10) |
| Increased | 28 (100%) |
| **Interleukin 6 (pg/ml, normal range 0-7) (N=11)** | 100.6 (51.51-919.5) |
| Increased | 11 (100%) |

Regarding the infection index, procalcitonin (PCT) rose above the normal range in 28 (100%) patients. 11 patients were tested for IL-6, most of whose index were above the normal range (Table 3). 32 patients had increased C-reactive protein (CRP).

Thirty-five (35, 97.22%) patients received antiviral treatment, including oseltamivir, ganciclovir, ribavirin or umifenovir hydrochloride. All patients were given antibiotic treatment (Table 2); 14 (38.89%) patients with a single antibiotic and 22 (61.11%) patients with combination therapy. 25 (69.44%) patients were also treated with glucocorticoids, and 20 (55.56%) patients treated with intravenous immunoglobulin therapy. Six patients have received aerosol inhalation treatment with a-IFN. Almost all the patients (35, 97.22%) received oxygen therapy, in which 19 (52.78%) patients used non-invasive ventilator mechanical ventilation and 9 (25%) patients used an invasive ventilator to assist ventilation.

All the cases had developed into Acute respiratory distress syndrome (ARDS). The median time from onset to ARDS was 11 days (Supplementary Figure 1). One (2.78%) patient presented with acute renal injury before ARDS. Electrolyte disturbance occurred in 16 patients (44.44%, Table 2). The median time from onset to death was 17 days (Figure 1).

**Figure 1.**
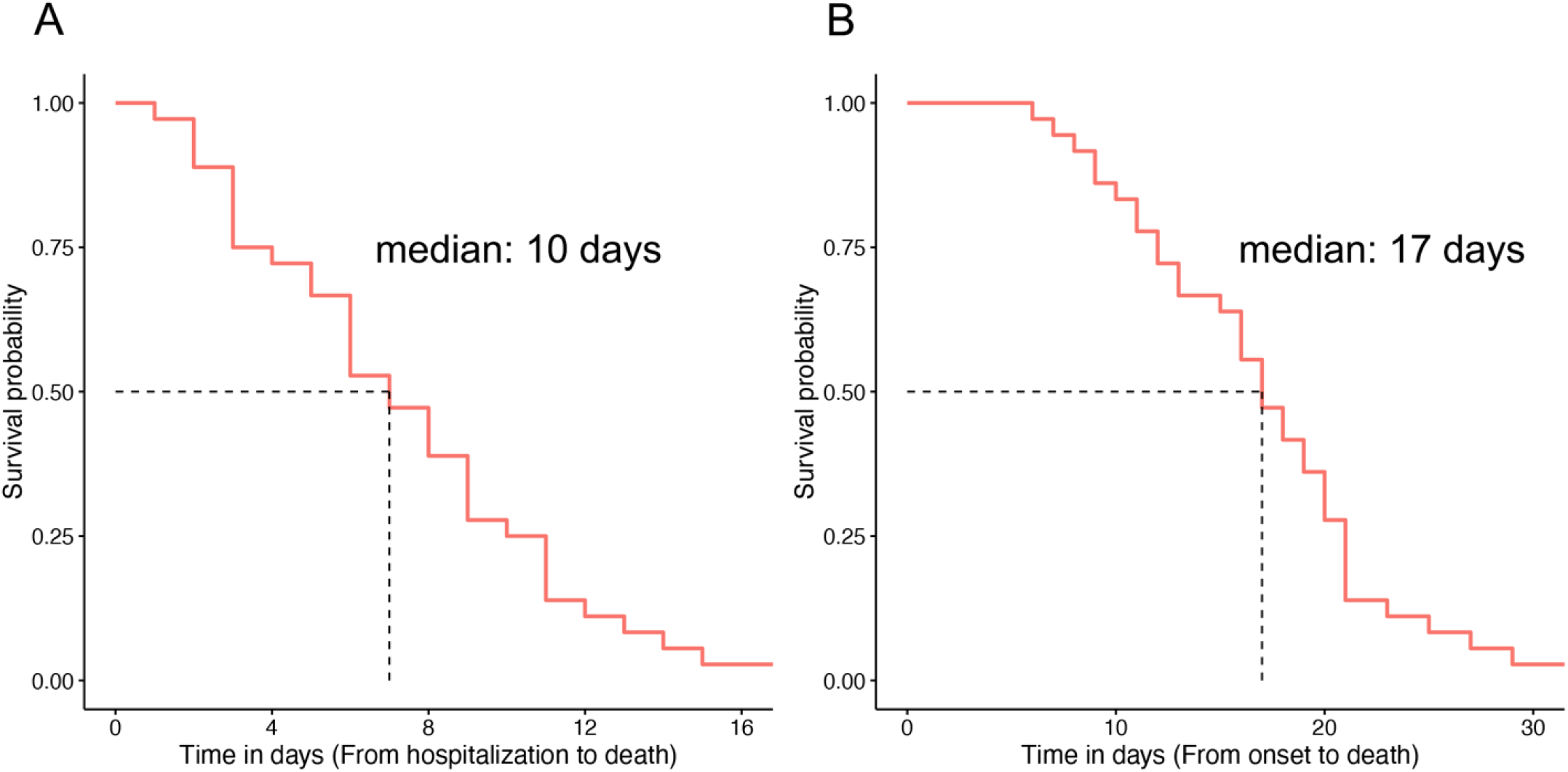
Survival curves of 36 non-survivors with COVID-19 in the Fifth Hospital of Wuhan. A. calculated from onset; B. calculated from hospitalization.

## Discussion

We reported on 36 patients who died from SARS-CoV-2 infection in a single hospital of Wuhan. All these patients had ARDS and 28 (77.78%) patients needed mechanical ventilation. Bilateral pneumonia was detected in most patients on admission. Previous studies have shown that elder patients and males who were confirmed with coronavirus infection are at higher risk of ARDS, ^10^ which was consistent with our study. Antibiotic treatment was used to prevent secondary infections since these patients were critically ill. Antiviral regimes were also used in most patients which we hoped may have roles but still need more data. However, there were no specific remedies for SARS-CoV-2 yet^11,12^. Therefore, most patients were treated with supportive treatment and intensive care. Unfortunately, due to restrictions of hospital’s conditions and the heavy burden of COVID-19 in Wuhan City, neither were we able to provide patients in need with advanced life support systems like Extracorporeal Membrane Oxygenation (ECMO), nor were able to transport all serious patients to superior hospital for better medical treatments in time. The median survival time of the patients in our study was only 2 weeks approximately, which was shorter compared to previous reports^13^. According to the study conducted by Yang et al., the mortality rate in critically ill patients with COVID-19 was higher than that in critically ill patients with SARS. To help patients who were critically ill survive from SARS-CoV-2 infection, advanced life support systems and other more intensive care are needed. In our cohort, non-survivors were hospitalized at 10 days after onset, and most of patients develop into ARDS within the first 3 days after admission (Supplementary Figure 1). This concentratedly reflected the current situation under the outbreak and was closely related to the run-up of medical resources. Early detection and intervention of patients are especially important which can delay the development from mild to severe cases. Encouragingly, more intensive care units (ICU) are being constructed, and more specialists and clinicians from different regions in China are assembling in medical institutions in Wuhan. Ultimately, the mortality of COVID-19 will decrease as the clinical capacity improves.

As reported in previous studies, patients who are male, elder or have a history of cerebrovascular disease are at higher risk of SARS-CoV-2 infection^2,14,15^. In our study, we found that the majority of coronavirus non-survivors were male patients, elder patients and patients with chronic diseases, which indicated that these patients might be also at increased risk of becoming critically ill or death. Cerebrovascular diseases were the most common comorbidities. Twenty-two (61.11%) non-survivors had cerebrovascular diseases in our report, and the percentage of cerebrovascular diseases among non-survivors was higher than that reported by Yang et al^13^. Besides, there were 4 (11.11%) non-survivors who had smoking history and 4 (11.11%) had preexisting COPD. According to previous studies on middle east respiratory syndrome coronavirus (MERS-CoV), smokers and COPD patients might be susceptible to MERS. Further, compared to non-smokers, smokers and COPD patients had a higher dipeptidyl peptidase IV (DPP4) expression, which was inversely correlated with lung function and diffusing capacity parameters^16^. Although the relationship between smoking history and susceptibility as well as worse outcomes in COVID-19 remains unclear, we cautioned that the prognosis of COVID-19 in patients with smoking history might be more severe.

In our cohort, lymphocytopenia occurred in more than 70% of patients at admission, which is a main laboratory feature in COVID-19 patients^15,17^. Lymphocytopenia have been identified in the critically ill patients with SARS-CoV and MERS infection^18,19^. As mentioned in previous studies, the severity of lymphocytopenia might indicate the severity of COVID-19, under the assumption of SARS-CoV-2 viral could attack and destroy the lymphocyte targetedly^2^. Further studies are warranted to confirm these findings. Increased levels of serum CRP, PCT, IL-6 were also found. It indicated the obvious inflammatory response among these patients. In addition, we noticed that the increased CRP, PCT concentration and decreased lymphocyte count from admission to death (Supplementary Figure 2), which may represent more prominent inflammation in severe patients. Therefore, intravenous glucocorticoids therapy, intravenous immunoglobulin therapy and interferon-alpha (α-IFN) aerosol inhalation were also used to restore homeostasis, without solid evidence. To our knowledge, there is still no specific medicine for COVID-19 till now. Clinical trials on promising regimens for COVID-19, such as remdesivir, lopinavir, and chloroquine phosphate are ongoing, which shed light on conquering the COVID-19 epidemic^12^.

This study has several limitations. First, only 36 dead cases were included. However, this is the largest cohort about the non-survivors of COVID-19 up to now. Second, some specific clinical information was insufficient, such as mechanical ventilation settings, oxygen concentration, and detailed medication history. Third, this is a retrospective, single-centered study. It may be limited to the hospital critical care resources. Further studies are still needed.

In conclusion, most non-survivors are older men with comorbidities (especially cardiovascular diseases). They finally progressed to ARDS. The median time from onset to ARDS was 11 days. The mean survival time in our cohort of COVID-19 non-survivors was about 2 weeks. Gradually decreased lymphocytes and increased inflammation biomarkers were found in these patients, and need to be monitored in the routine treatment. Early detection and intervention of patients are especially important which can delay the development from mild to severe cases.

## Data Availability

The data will be made available to others on reasonable requests to the corresponding author. Deidentified participant data will be provided after approval from the corresponding author and the Fifth Hospital of Wuhan.

## Contributors

YH, RY,YX collected the epidemiological and clinical data. YH summarised all data. YH, RY, YX, and PG drafted the manuscript. YH and PG revised the final manuscript.

## Declaration of interests

We declare no competing interests.

## Acknowledgments

We thank all patients and their families involved in the study. We thank all the doctors and nurses of our hosiptal (see Supplementary Appendix) for their efforts in taking care of these patients. We especially thank all the doctors and nurses of Jiangxi Support Team.

Supplementary Appendix of the staff who took care of the 36 patients in this article:

1. Song Hu, Zhixiong Liang, Yousheng Yang, Xuejun Xiao, Zhi Zhang, Zekun Hu, Yan Peng, Department of Critical Care Medicine, the Fifth Hospital of Wuhan, China
2. Guizhong Xiong, Department of Stomatology, the Fifth Hospital of Wuhan, China
3. Xiaohong Lv, Dongchu Wang, Juping Han, Xiaolan Zheng, Chenglu Tang, Weibo Wang, Lihua Shao, Xiaoxia Li, Qian Wu, Longyan Chen, Department of Gastroenterology, the Fifth Hospital of Wuhan, China
4. Gang Zheng, Wenjing Chai, Feixiang Chen, Liuheng Deng, Department of General Surgery, the Fifth Hospital of Wuhan, China
5. Tao Xiong, Mei Zhang, Qiang Li, Xian Zhang, Quan Zhou, Shuchun Ou, Renlin Hu, Department of Neurology, the Fifth Hospital of Wuhan, China
6. Jingfen Lu, Department of Endocrinology, the Fifth Hospital of Wuhan, China
7. Dan Ma, Huijun Fan, Zilong Gong, Yue Wu, Qinghong Shi, Dan Zhang, Dan Liu, Qiong Wang, Qian Li, Wanxian Yu, Department of Respiratory, the Fifth Hospital of Wuhan, China
8. Xiulan Peng, Department of Oncology, the Fifth Hospital of Wuhan, China
9. Hanping Chen, Zecheng Wei, Ruiguang Weng, Biao Dong, Department of Neurosurgery, the Fifth Hospital of Wuhan, China
10. Yang Liu, Minghui Li, Lei Wu, Hongjun Xu, Lei Yan, Liang Huang, Fangzhou Chen, Department of Orthopedics, the Fifth Hospital of Wuhan, China
11. Hao Li, Xuechuan Liu, Mi Zhang, Jingtao Li, Department of Urology Surgery, the Fifth Hospital of Wuhan, China
12. Mingqiao Ding, Weichen Zhang, Yaming Hao, Department of traditional Chinese medicine, the Fifth Hospital of Wuhan, China

## Supplementary materials

Supplementary Figure 1. The disease course of 36 non-survivors. A. Time in days from onset to hospitalization; B. Time in days from onset to ARDS; C. Time in days from ARDS to death.

Supplementary Figure 2. Decreased lymphocyte count in non-survivors (between admission and last laboratory test before death).

## Notes

### Competing Interest Statement

The authors have declared no competing interest.

